# The phenomenology of psilocybin’s experience mediates subsequent persistent psychological effects independently of sex, previous experience or setting

**DOI:** 10.1101/2024.08.26.24311611

**Authors:** Tereza Klučková, Marek Nikolič, Filip Tylš, Vojtěch Viktorin, Čestmír Vejmola, Michaela Viktorinová, Anna Bravermanová, Renáta Androvičová, Veronika Andrashko, Jakub Korčák, Peter Zach, Kateřina Hájková, Martin Kuchař, Marie Balíková, Martin Brunovský, Jiří Horáček, Tomáš Páleníček

## Abstract

**Background:** Recent studies have intensively explored the potential antidepressant effects of psilocybin. However, important variables such as previous experience, repeated administration, setting and sex remain underexplored. This study describes the acute psilocybin experience and long-term effects in a small sample of healthy individuals.

**Methods:** In a double-blind, placebo-controlled, cross-over study, 40 healthy participants (20 females, mean age 38, sd 8) received two doses of psilocybin 0.26 mg/kg per os at least 56 days apart (mean 354 days) in two study arms (EEG and fMRI). Near half of participants had experience with psychedelics. The Altered State of Consciousness Scale (ASC) and a visual analogue scale (VAS) on emotional valence of the phenomenology assessed acute phenomenology. The Persisting Effects Questionnaire (PEQ) assessed long-term effects. Venous blood samples were taken to measure serum psilocin levels.

**Results:** All results were independent of previous experience, sex, EEG or fMRI arm/setting. Acute psychedelic effects were of moderate intensity on ASC. The VAS showed mostly pleasant and fluctuating, and only one unpleasant only experience. All experiences resolved in a positive or neutral state at the end of the session. Psilocybin induced sustained positive effects on all domains of the PEQ, with negligible negative effects. Oceanic Boundlessness and Visual Restructuralization were associated with positive effects on PEQ. Contrary to expectations, Dread of Ego Dissolution, typically associated with fearful experiences, was not associated with PEQ negative outcomes. The type of experience (pleasant or mixed) did not correlate with the intensity or direction of the lasting effect; however, peak experiences culminating in a positive mood were associated with positive long-term effects.

**Conclusion:** In our sample repeated administration of psilocybin to healthy volunteers, induces positive, lasting effects. This underscores the psychological safety of psilocybin in a laboratory setting and supports its repeated use in clinical trials. In particular, challenging or anxiety-provoking experiences in controlled environments did not lead to adverse long-term outcomes.

Clinical trial registration: EudraCT 2012-004579-37, https://www.clinicaltrialsregister.eu/ctr-search/trial/2012-004579-37/CZ.

## 2. Introduction

Psilocybin, a natural serotonergic psychedelic of the tryptamine family, has recently gained robust evidence as a potential antidepressant ^1–4^. Other studies also suggest its benefits in terms of positive, sustained changes in personal well-being and attitudes towards self and others in healthy subjects ^5–7^. Unlike classical antidepressants, psilocybin’s antidepressant effects occur almost immediately after a sufficiently potent dose. The onset of action is comparable to that of the dissociative anaesthetic, NMDA antagonist, ketamine, which is now widely used off-label as an antidepressant in patients ^8,9^. Because of these rapid effects, psilocybin, ketamine and other psychedelic drugs have been combined into a new class of so-called rapid-onset antidepressants ^10–12^.

One of the key questions regarding the neurobiology of the antidepressant effects of psilocybin and other psychedelics is the extent to which the psychedelic experience drives the antidepressant effect. While there are some indicators of the importance of psychedelic or psychotomimetic effects, or the peak and mystical type of experience, for the long-term positive outcomes of psilocybin and ketamine in both healthy and clinical populations ^3,13–23^, other research, with ketamine and esketamine, suggests that similar effects can be achieved only pharmacologically, bypassing the psychedelic state ^24–29^. Subsequent critical questions concern the consistency of these positive long-term effects with repeated administrations and their influence on phenomenology. In addition, the influence of other variables, such as the role of sex, set and setting, warrant investigation.

The psilocybin-induced experience is characterised by perceptual, emotional, and cognitive alterations and a subjective loss of identity, described as ego dissolution. The positive nature of ego dissolution in terms of being a predictor of beneficial psychological effects is often referred to as a peak ^30^, mystical ^31–33^ or enlightening experience ^34^. It is thought to be one of the most important factors in determining long-term outcomes following psychedelic treatment ^35,36^. These acute effects of psilocybin are typically assessed using Altered State of Consciousness (ASC) scales, the Mystical Experience Questionnaire (MEQ) and/or visual analogue scales ^37^. The intensity and positive valence of the psychedelic experience when culminating in a ’peak’ experience seems to predict beneficial long-term outcomes ^21,38^, while intensive anxiety seems to be associated with the opposite ^21^. The proposed mediators of the effect of peak experiences on long-term outcomes are the increase in psychological flexibility ^39,40^, awe ^41^ and changes in brain criticality ^42,43^. Previous findings in healthy subjects have shown that repeated psychedelic experiences result in less acute effects ^44,45^ and found no effect of sex. However, in contrast to acute effects, there is no evidence on the role of these factors on long-term outcome.

Given these considerations, the primary objective of this study is to elucidate the relationship between acute phenomenological experiences and potential antidepressant-like effects in healthy subjects. Specifically, the study aims to examine the correlation between phenomenology and the enduring effects of the psilocybin experience, accounting for variables such as prior experience, repeated administration, sex. Unlike in patient populations, assessing antidepressant-like effects in healthy individuals requires an emphasis on more nuanced alterations, including overall well-being and mood shifts, which may not attain a level of pathological significance. Such persistent changes are captured by the Persisting Effects Questionnaire (PEQ) ^5,6^, which effectively measures attitudes to self, life, mood, relationships, behaviour and spirituality. The PEQ was constructed on the basis of previous research that reported positively valued psilocybin experiences in healthy volunteers ^46^, which had been shown to have personal significance even after 25 years ^47^. The PEQ also showed optimal convergent validity with the phenomenology of the experience as assessed by the Altered State of Consciousness Scale (ASCs or APZ) and its renewed five-dimensional version of the ASCs (5DASCs) ^48–50^.

Thus, here we tested the effects of the acute phenomenology of the psilocybin experience, assessed by ASCs and visual analogue scales describing the course and directionality of the emotional valence of the experience, on the long-term effects on mood, well-being and spirituality, assessed by PEQ, in healthy subjects, controlled for previous experience with psilocybin, treatment order and sex. As the data presented here come from a larger study that is also evaluating the neurobiology of psilocybin’s effects from an EEG and fMRI perspective, we have collected data from participants exposed to psilocybin repeatedly in two different settings (EEG and fMRI). Based on the literature, we hypothesize that the psilocybin-induced psychedelic experience will drive long-term positive outcomes, while the above variables will be treated as exploratory measures.

## 3. Methods

### 3.1. Study approval

The study was approved by the Ethical Committee of the National Institute of Mental Health (NIMH-CZ), by the State Institute for Drug Control and as a clinical trial registered under the EudraCT No. 2012-004579-37.

### 3.2. Experimental design

Data were collected in two consecutive arms in which subjects underwent a battery of questionnaire-based, phenomenological, neurocognitive and EEG or fMRI measurements. While the first arm was designed to collect primarily EEG data (EEG arm), the second arm was designed to collect primarily fMRI or simultaneous EEG/fMRI measurements.) As the study allowed for the enrolment of inexperienced subjects, the EEG arm always preceded the fMRI arm, as we agreed that the procedures associated with EEG were less stressful, so that we did not expose inexperienced subjects directly to the demanding environment of fMRI. In each arm, subjects received both treatments, psilocybin and placebo, in a double-blind crossover design. The minimum interval between sessions was 28 days. See Figures 1.A & B for details.

**Figure 1.**
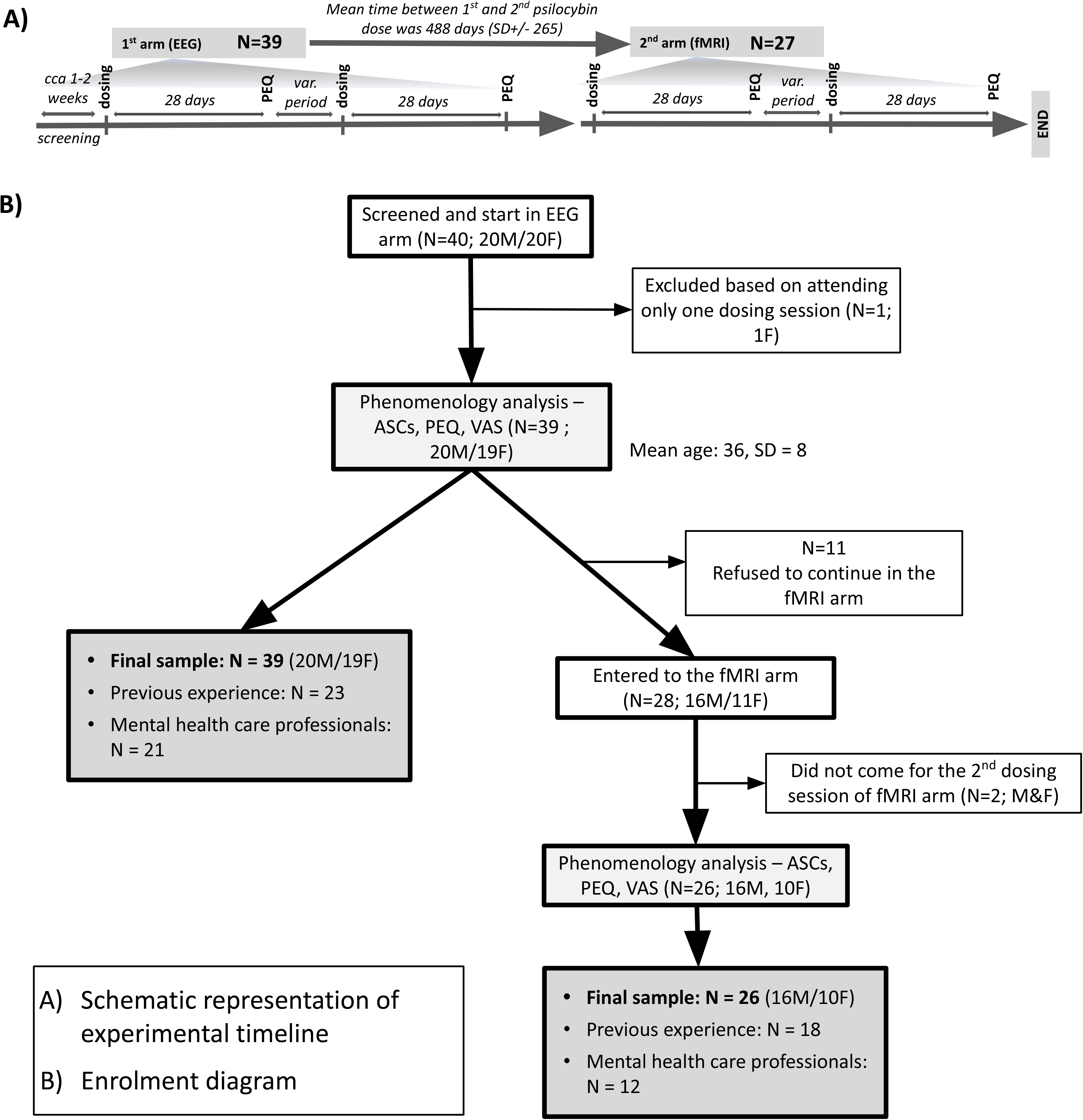
A) experimental timeline, **B)** study flow chart. PEQ = Persisting Effects Questionnaire, VAS = Visual Analog Scale, ASCs = Altered state of Consciousness Scale

Throughout the study, each participant was accompanied by a fixed pair of sitters consisting of one man and one woman, one of whom was a psychiatrist. Prior to dosing, details of the study procedures and drug effects were discussed during a preparatory session. Each dosing session began with a brief clinical examination to check inclusion/exclusion criteria. This included a somatic examination, questions about a current living situation, blood pressure and heart rate, and a negative result on a breathalyser test and a urine drug screen. An intravenous cannula was inserted for blood sampling and an EEG cap was placed on the participant’s head before the session. The 6-8 hour session, starting at 7-8 am, took place in a decorated EEG experimental room, where the subjects spent most of the experiment. In the case of the fMRI arm, subjects went to an MRI scanner in a neighbouring laboratory on three occasions (baseline, 90 and 240 min after dosing) for a 45-60 min scanning session. The treatments used were oral psilocybin 0.26 mg/kg and placebo (Tritici Amylum). The formulations - capsules containing either 5 or 1 mg of psilocybin - were prepared in a pharmacy at IKEM (Institute of Clinical and Experimental Medicine in Prague). The dose of psilocybin was adjusted by combining capsules containing 1 and 5 mg, increasing or decreasing by 1 mg per 5 kg of body weight, with a 75 kg person being treated with 20 mg of psilocybin. In the case of placebo, the number of capsules was identical. The average dose used in the study was 18.6 mg (15 - 24 mg). Subjects took the capsules on an empty stomach with 200 ml of water. The study nurse was also present throughout the session in a neighbouring room. The women’s sessions were scheduled outside the menstrual phase of the female cycle to reduce interindividual variation due to the effect of menstruation and to avoid potential differences associated with different phases of the cycle ^51^. After the acute effects had subsided, participants completed questionnaires assessing the quality of the experience. At the follow-up, 28 days after dosing, participants were invited to complete a questionnaire on the persistent effects attributed to their last session.

### 3.3. Participants

Recruitment was carried out using a peer-to-peer snowball sampling method. It started with an initial pre-screening by telephone interview for key exclusion criteria (see Appendix 1 for details). The study was designed to balance the whole group in terms of sex (M/F) and previous experience (50% naive subjects). Although this was not the primary aim, the snowballing method resulted in a near-balanced sample in terms of mental health and non-mental health education/work status. Eligible participants were invited to a face-to-face interview. Participants were informed of the study design, the effects of psilocybin, safety concerns, and that they were free to withdraw from the study at any time. Participants were given space to ask questions. Informed consent was obtained. Participants were screened using the Minnesota Multiphasic Personality Inventory (MMPI-2) ^52^ and the Mini-International Neuropsychiatric Interview (MINI v 5.0) ^53^ to rule out psychopathology and were physically examined to rule out somatic problems ^54^. 40 participants were included in the study. See Figure 1.B for the exclusion process and the final sample. 26 subjects participated in both the EEG and fMRI arms, as 14 subjects did not continue in the fMRI part of the study. The main reason for not continuing was that they were afraid of the fMRI setting and tasks to be disturbing and a long time frame between the EEG and fMRI arm. Details of the enrolment process and sample characteristics are shown in the flowchart in **Fig. 1** and in the **Tab. 1**.

**Table 1:**
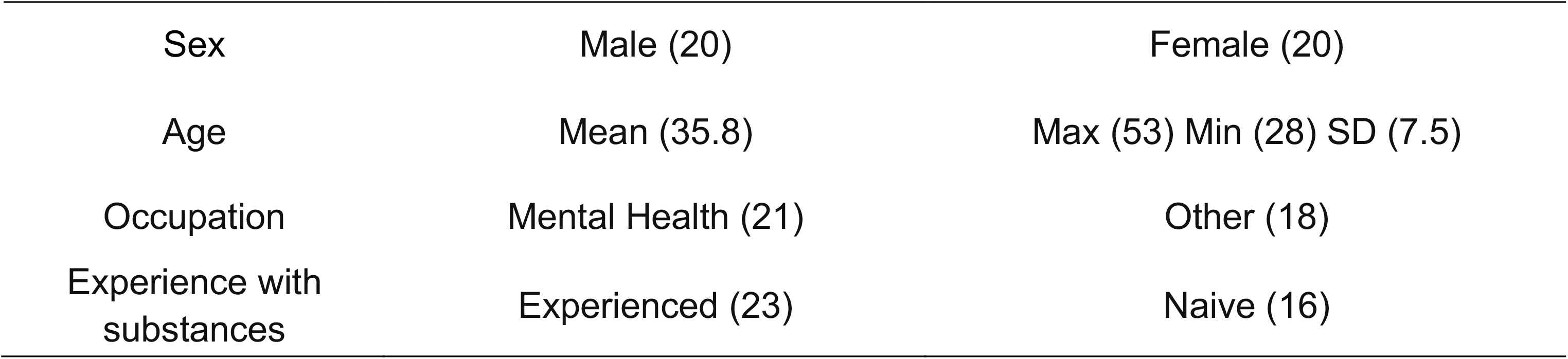
Sample description N = 40

|  |  |  |
| --- | --- | --- |
| Sex | Male (20) | Female (20) |
| Age | Mean (35.8) | Max (53) Min (28) SD (7.5) |
| Occupation | Mental Health (21) | Other (18) |
| Experience with substances | Experienced (23) | Naive (16) |

### 3.4. Altered State of Consciousness scale (ASC)

Immediately after the session, the participants completed the Czech version of the ASC questionnaire (in the original: Aussergewöhnliche Psychische Zustände APZ) ^48^. It consists of 72 questions assessing the subjective acute effects of the altered state of consciousness in 3 subscales and 1 total scale including all questions: Oceanic Boundlessness (OBN), Dread of Ego Dissolution (DED) and Visionary Restructuralisation (VRS), and the General - Altered States of Consciousness (G-ASC).

The OBN scale refers to a deeply positive experience of depersonalisation and oneness. High scores indicate a mystical experience of oneness. The DED scale refers to a negative experience of depersonalisation, loss of control and increased anxiety. High scores indicate a difficult and probably unpleasant nature of the experience, which, if it persists throughout the experience, can be referred to as a bad trip in an uncontrolled environment. The VRS scale refers to perceptual alterations and includes symptoms such as illusions, (pseudo) hallucinations, synesthesia, typically but not limited to the visual domain. The secondary scale G-ASC consists of all 72 items of the questionnaire and can be interpreted as a general measure of altered consciousness.

### 3.5. Persisting Effects Questionnaire (PEQ)

The PEQ ^5,6^ consists of 140 questions in 6 categories, which are subsequently divided into positive and negative categories: attitudes to self, attitudes to life, mood changes, altruistic social impact, behavioural changes, and spirituality; and 3 additional questions on personal meaning, spiritual meaning and change in life satisfaction. Most items are rated on a 6-point scale (0 = not at all; 1 = so slight cannot decide; 2 = slight; 3 = moderate; 4 = strong; 5=extreme, more than ever before in your life and stronger than 4). The three additional questions are: (1) How personally meaningful was the experience? (Rated from 1 to 8, with 1 = no more than routine, everyday experiences; 7 = among the five most meaningful experiences of my life; and 8 = the single most meaningful experience of my life); (2) Indicate the degree to which the experience was spiritually significant to you? (Rated from 1 to 6, with 1=not at all; 5=among the five most spiritually significant experiences of my life; 6 = the single most spiritually significant experience of my life); (3) Do you believe that the experience and your contemplation of that experience have led to change in your current sense of personal well-being or life satisfaction? (Rated from + 3 to - 3, with + 3 = increased very much to – 3 = decreased very much)

### 3.6. Visual Analogue Scale (VAS)

At the end of the dosing session, subjects were asked to plot the course of the pleasant/unpleasant emotional valence of their experience as a continuous curve over 7 hours of experience on a 2D graph (time from 0 to 7 in hours was on the X-axis; Y-axis = pleasant/unpleasant experience, VAS range 10 to -10), according to the following task "Plot a continuous graph of how your intoxication progressed in terms of pleasant and unpleasant (anxious) experiences ^55^. Use the highest values only for ecstatic states or states of opposite extreme polarity". The plotted forms were then digitised using the freely available tool Webplotdigitizer with the algorithm "X step w/ Interpolation" with a step size of 0.1 and a smoothing of ΔX of 200%.

### 3.7. Exploratory analyses

We examined the associations between the VAS scale and persisting effects. Based on a closer look at our data, we have tested whether subjects with ’positive only’ and ’positive and negative’ experiences would differ in their persisting effects (**Fig. 4**). Subsequently, we also evaluated whether the peak of the experience or the last point of the experience or their combination would be associated with the long-term outcome ^21^, see **Fig. 5B in online supplemental**.

### 3.8. Psilocin serum levels

Blood samples were taken before the experiment and at 1, 2, 3 and 6 hours after dosing. During the course of the study, due to external conditions, we had to change the laboratory performing the measurements of psilocin serum levels, switching from earlier measurements performed by the GC-MS method (first 20 subjects in the EEG arm) ^54^ to the remaining measurements performed by LC-MS ^56^.

### 3.9. Data processing and statistical analyses

Descriptive statistics were calculated. Independent variables were sex, previous experience, and placebo/psilocybin order. Dependent variables were ASC and PEQ scores. Normality of the data was assessed visually (QQ plots) and statistically (Shapiro-Wilk test). PEQ and ASCs scores were normalised to percentages to facilitate visual understanding. Data from the EEG and fMRI arms were treated as separate samples due to an average of 1 year between sessions and attrition. Data were tested for equal variances and the difference between the first and second psilocybin session was compared.

Correlations between ASC and PEQ subscales were calculated and corrected using the Holms-Bonferroni procedure.

VAS data were integrated into the total area under the curve (AUC) and the total positive area under the curve (only values above zero; pAUC). The relative index of positive experience (RIPE) was calculated as the ratio of AUC to pAUC. The RIPE index was used to divide the sample into those who had only positive experience and those who had both positive and negative experience. This new factor was then used in a secondary analysis of ASC and PEQ. A two-sample t-test was used to detect a difference. P-values were not corrected.

In an exploratory analysis, the average of the highest VAS score and a score at 6 hours was calculated for each participant (peak-end rule) ^57^ and correlated with the dependent variables.

Statistical analysis was performed using RStudio software for iOS, version 1.4.1743 (RStudio, Inc., 2009-2022).

## 4. Results

### 4.1. Sample characteristics and psilocin serum levels

Sample is described in **Fig. 1**, **Tab. 1**. Psilocin serum levels are seen in **Fig. 5A,C in online supplemental**.

### 4.2. Altered state of consciousness scale (ASCs)

There was a significant difference between the psilocybin and placebo conditions in both the EEG and fMRI arms (**Fig 2A,C**). T-tests showed a difference on each subscale (p < 0.001 values adjusted for multiple comparisons). There were no significant differences in between-subjects variables (sex, experience, occupation; results of F-statistics are shown in **Tab. 2 in online supplemental**). There was no significant difference between the intensity of the first and second psilocybin experiences.

**Figure 2:**
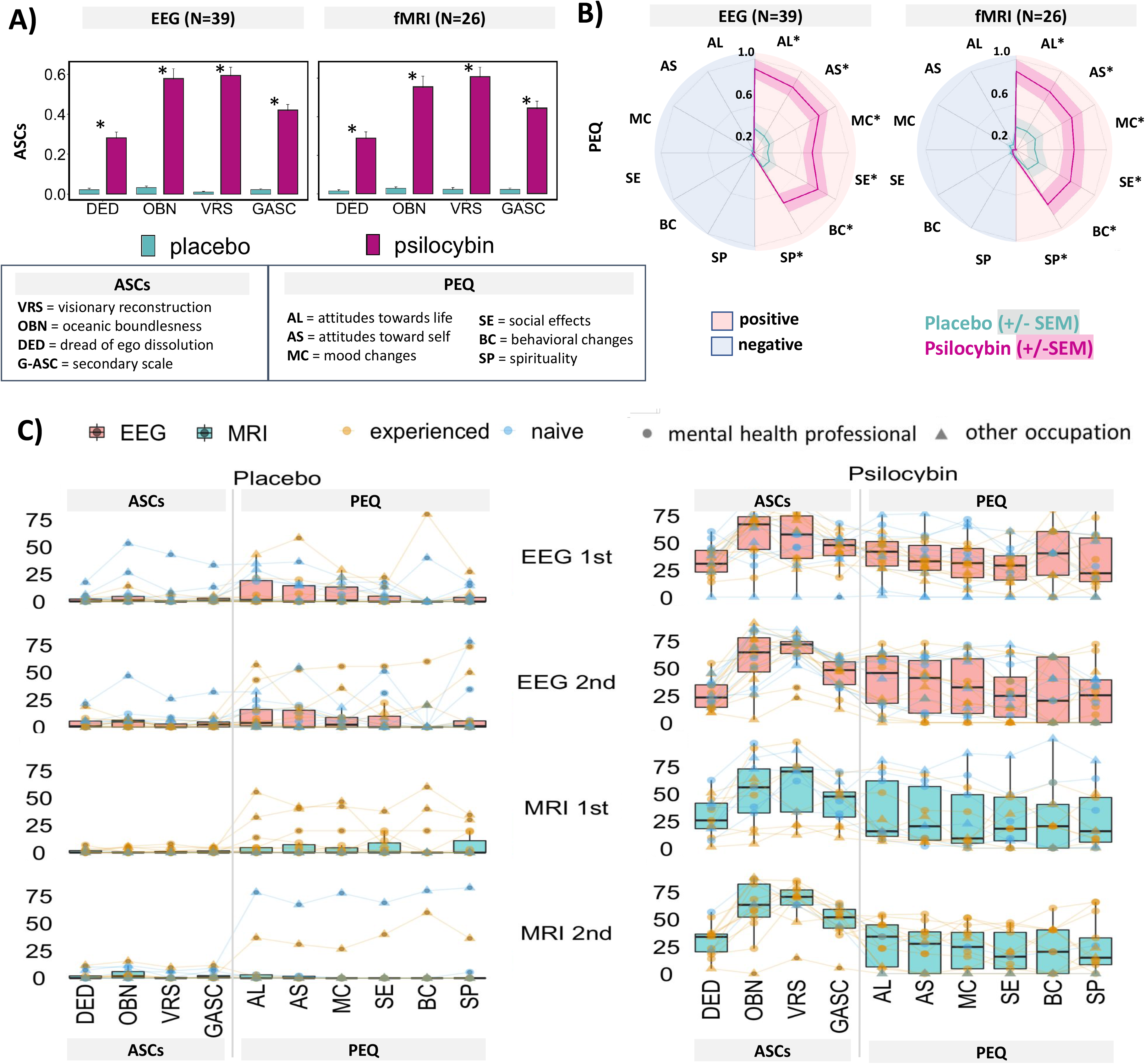
A) ASCs scores: Overall differences between psilocybin and placebo. Psilocybin induced significant increase in all domains of ASCs compared to placebo, there were no significant differences between EEG and fMRI arms. *\* indicates P ˂ 0.05 for psilocybin versus placebo (corrected)* **B)** PEQ scores: Overall differences between normalized PEQ scores between psilocybin and placebo. Negative effects are depicted in blue in the left half of the circle, positive subscales in red and right half. Psilocybin as well as placebo had negligible negative effects and induced primarily positive persisting effects. Psilocybin was significantly more potent compared to placebo in all subscales. *\* indicates P ˂ 0.05 for psilocybin versus placebo (corrected), the shaded strip around the line corresponds to +/-SEM*. **C)** Shows in details the differences between ASCs and PEQ in the two study arms (EEG and fMRI) visualizing the effect of order, whether subjects were previously experienced or not and whether they were mental health professionals or other professions. The graph also depicts individual flow of subjective experience to persisting effects and clearly shows the trends in placebo responders and psilocybin non-responders.

### 4.3. Persisting effects scale (PEQ)

We found a significant difference between the psilocybin and placebo conditions for all positive subscales (**Fig. 2B,C**; p<0.001, p. values adjusted). Most subjects did not experience any negative effects, and negative subscales were close to 0, so further statistical analysis in this direction was not pursued. ANOVA revealed no significant differences in the between- subjects variables (results of F-statistics in **Tab. 3 in online supplemental**). There was no significant difference between the first and second experience.

13 out of 39 participants (33%) rated the psilocybin experience as one of the top 5 most meaningful experiences of their lives. The experience increased the current sense of personal well-being or life satisfaction moderately for 14 participants (36%) and very much for 10 participants (26%). 6 participants (15%) rated the psilocybin experience as the single most spiritually significant experience of their lives, and a further 8 participants (21%) rated it among the top five most spiritually significant experiences.

### 4.4. Correlation between ASCs and PEQ: Positive

**Figure 3** shows the correlation matrix for EEG and fMRI. In the EEG arm, all ASCs subscales were positively correlated with long-term positive PEQ outcomes. Except for DED, all correlations survived corrections. In the fMRI arm, the effects were less pronounced and none survived corrections.

**Figure 3.**
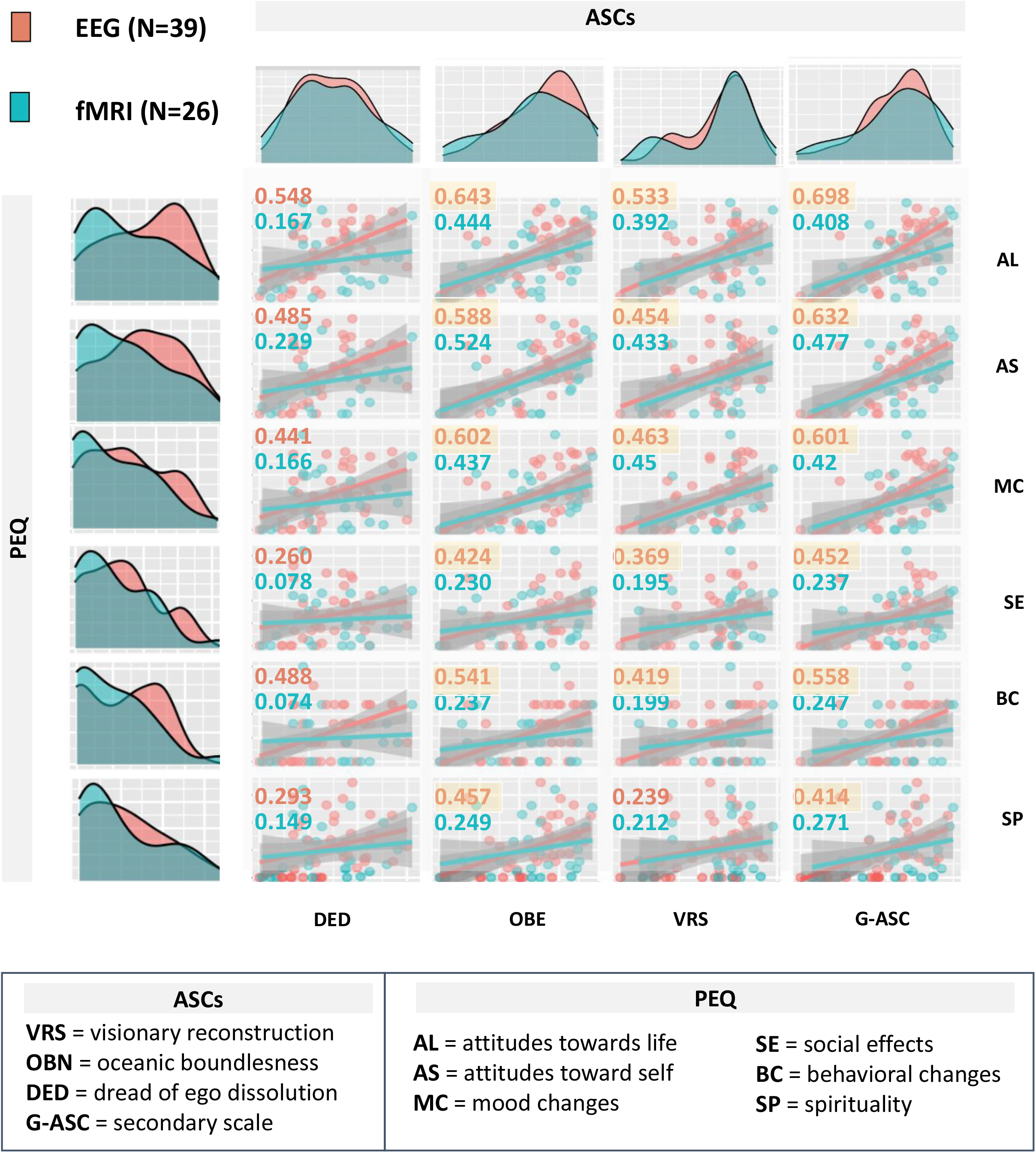
Distributions, Scatterplots, and correlations between ASCs and PEQ scores. EEG (red) and fMRI (blue) branch. Score distributions show difference between EEG and fMRI on ASCs and PEQ scores. In yellow are highlighted correlation values that survived the Bonferonni-Holms correction.

**Figure 4.**
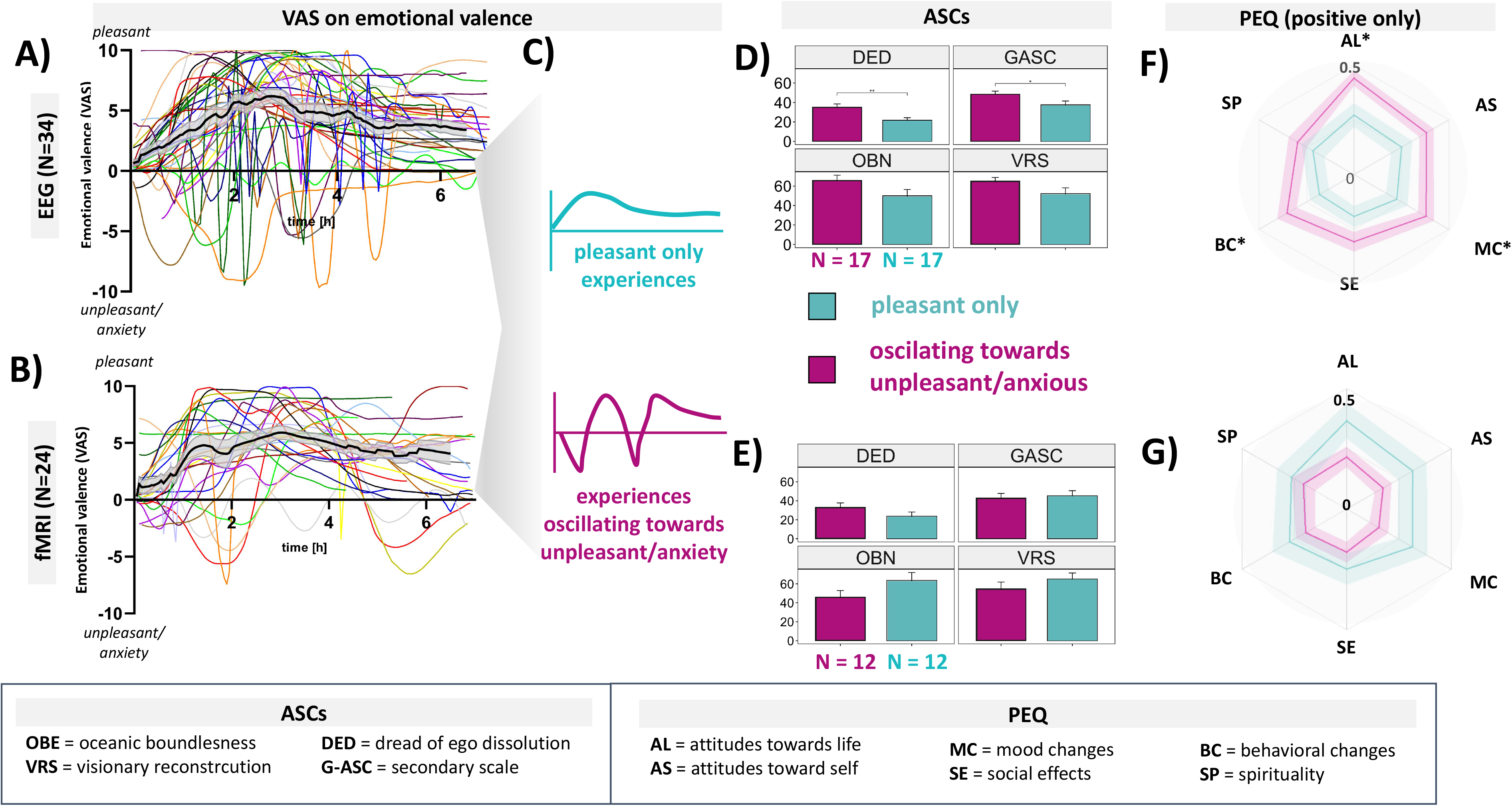
<u>VAS scale following psilocybin experience:</u> The course of emotional valence across time in EEG branch **(A)** and fMRI branch **(B)**. Each colored line represents the course of one individual experience, the blackline with grey strip around represents Mean (+/-SEM). While individual experiences could oscillate between pleasant and unpleasant/anxious experience several times, the means, however, refer to overall pleasant experience. Almost all individual experiences ended at either 0 or in the emotionally pleasant state at the end of observational period. In order to reveal whether participants that experience pleasant only experiences would have different phenomenology and long-term outcome compared to subjects who had experiences oscillating towards unpleasant/anxious pole we have split the sample into these two subgroups in each arm **(C)** and compared the ASCs **(D, E)** and PEQ **(F, G)** within these. In the EEG arm **(F)**, participants who had both positive and negative experience scored higher in ASCs on DED, and GASC but not on VRS while in the fMRI arm (**G)** subgroups did not differ significantly. Similarly, significant difference within PEQ was present on the AL, MC and BC subscales in EEG arm **(F)** while no difference was observed in the fMRI arm (G). *\* indicates P ˂ 0.05 for comparisons of the pleasant vs unpleasant/anxious groups (p. val. uncorrected)*

**Figure 5.**
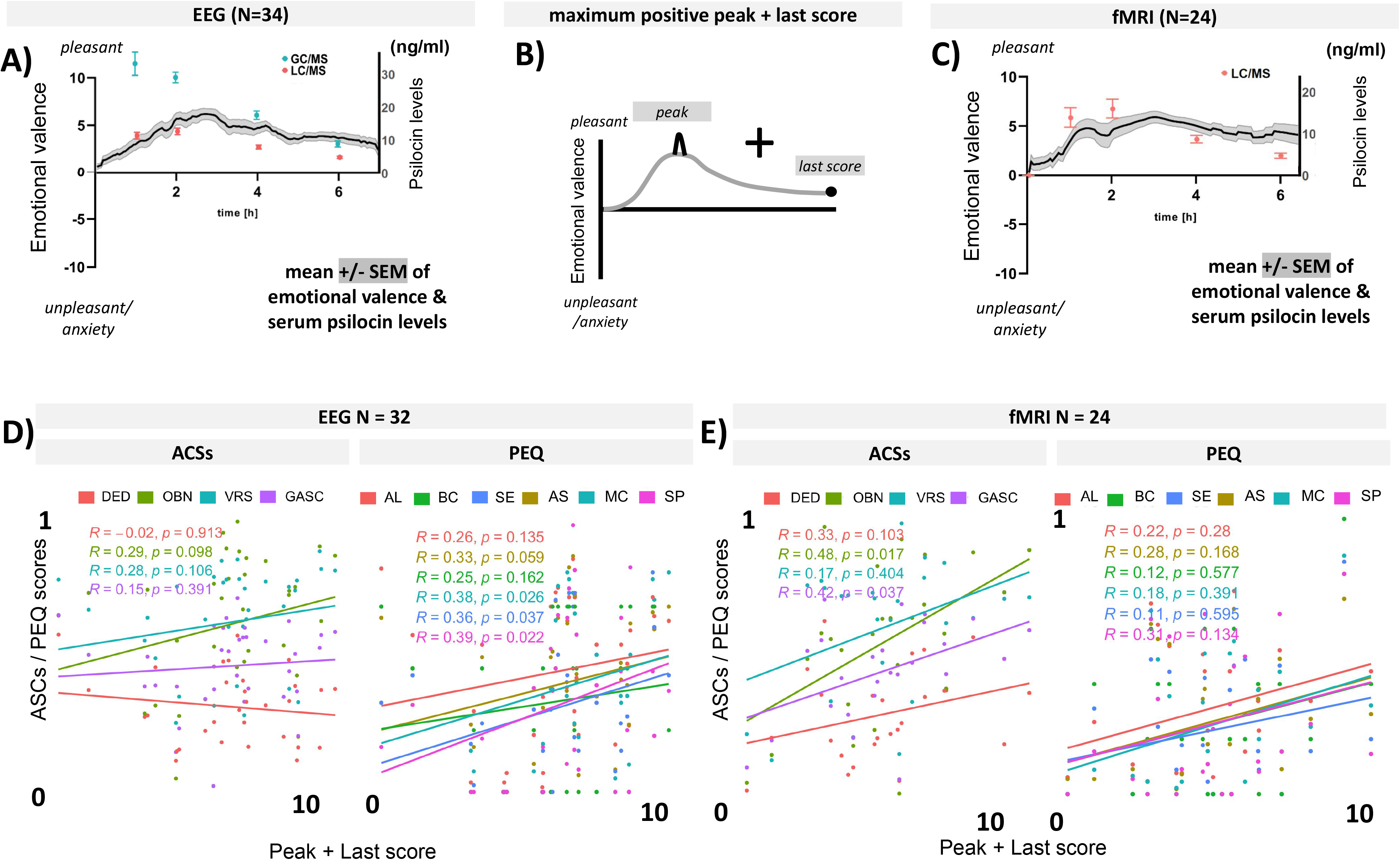
(A, C) visualization of VAS scale on emotional valence with psilocin levels measured by (GC/MS and/or LC/MS) in EEG and fMRI arms. **(B)** schematic visualization of peak + last point hypothesis. Below are correlations of ASCs and PEG subscales with the „maximum positive peak + last score“ within EEG arm **(D)** and fMRI arm **(E).** *Shown P values are uncorrected)*

### 4.5. Visual Analogue Scale (VAS)

**Figures 4A and 4B** show the individual emotional valence as well as the mean and standard error. There were three types of experience: 1) positive only (EEG: n = 17; fMRI: n = 13), 2) oscillating between positive and negative (EEG: n = 17; fMRI: n = 13) and 3) negative only (EEG: n = 1). At the end of the sessions, mean experiences converged towards a positive emotional state, at the individual level the majority ended in positive or returned to baseline, and three subjects in the fMRI arm ended in a slightly negative emotional state.

### 4.6. Additional exploratory analysis

The relative index of positive experience (RIPE) was calculated from the VAS scale scores. For example, a score of 1 can be approximated as 100% of the experience being positive, whereas 0.83 can be approximated as 83% of the experience being positive. RIPE allowed us to divide the sample into RIPE equivalents above and below a given threshold. We chose an arbitrary threshold of 0.98, which conveniently divides the sample into two of similar N.

By dividing the sample in this way **(Fig. 4C)**, the ASC and PEQ subscales were analysed. For ASC in the EEG arm, DED, OBN and GASC were significantly higher in subjects who had both positive and negative experiences compared to those who had only positive experiences. For the EEG arm, participants with both positive and negative experiences scored higher on the PEQ for positive self-attitudes, positive mood changes, and positive behavioural changes. However, none of these effects were present in the fMRI arm (see **Fig. 4D, E, F and G**). P-values were not corrected.

Correlations between peak/last scores showed mild positive correlations (uncorrected) for a number of subscales on the PEQ **(Fig. 5B, D, E in online supplemental)**.

### 4.7. Responders and non-responders

There were 4 non-responders to psilocybin and 6 placebo responders, non-response to psilocybin, and response to placebo, was defined as the ASC score being similar to the mean of the opposite group and/or scores in the 1st or 3rd quartile. One non-responder had very low serum levels of psilocybin compared to a comparable subject (women around 50 years old, with the same dose of 18 mg of psilocybin). The placebo responders show two types of experience. First, change in a state of consciousness for up to an hour and half, followed by a return to normality. This may be explained by the effect of expectancy and the environment, followed by a functional unblinding, and this experience is seen only in the first session of each arm, where participants were uncertain about condition assignment. The second type of experience, which may be called the true placebo response, subjects experienced psychedelic effects throughout the session (see **Table 4. in online supplemental**).

At the time of the study design, the nocebo effect was not yet an understood concept, and from available data, we cannot infer the occurrence of a nocebo effect. This may be because in any case, participants knew they will receive psilocybin in one of the conditions.

## 5. Discussion

This study has demonstrated that the overall intensity of the psilocybin experience, at a dose of 0.26 mg/kg, is of moderate level, encompassing the entire range of effects/dimensions as characterized by the Altered State of Consciousness Scale (ASCs), with notably lower levels of Dread of Ego Dissolution (DED) compared to Oceanic Boundlessness (OBN) and Visionary Reconstruction (VRS) subscales. Utilizing visual analogue scales, we observed participants with exclusively positive effects, those with fluctuating experiences including challenging or anxious moments, and only one individual who had a solely difficult experience on one occasion. Predominantly, participants ended their sessions in either a neutral or positive emotional state, with only three reporting a marginally negative mood at the end. The experiences were predominantly rated as having lasting positive or neutral impacts on well-being, with minimal negative effects that did not reach clinical or statistical significance. Our findings indicate no significant influence of sex, previous psilocybin exposure, or setting (EEG vs. fMRI) on the ASCs and PEQ outcomes. Intriguingly, all ASC dimensions, including DED, correlated positively with persistent positive effects, suggesting that the overall intensity of the experience is a key driver of this relationship. Exploratory analyses further imply that the nature of the peak experience, in conjunction with the emotional state at session’s end, may serve as an indicator for predicting the long-term impact of individual psychedelic experiences.

### 5.1. ASCs, VAS, PEQ

The overall effects on ASC and PEQ are in line with previously published data on comparable items ^5,6,21,38,45,58^ and correspond to lower medium / optimal doses of psilocybin ^59–61^, with about half of the subjects having a full psychedelic experience. The added value of our study is that we found no difference on the ASC and PEQ between naive and experienced participants, nor between the first and second psilocybin administration. Furthermore, there were no differences in how many of the subjects in our sample rated the experience as one of the most significant in their lives, regardless of their previous experiences. This is partly in contrast to ^44,45^, where psychedelic naive subjects reported stronger effects on Disembodiment, Visual Restructuralisation and Changed Meaning of Percepts of the 5D- ASCs. One explanation for our observations could be that all participants may have perceived the experience as equally intense due to the overall novelty of the setting, which would mask their previous experience. Also, most of the experienced subjects had used psilocybin in a recreational setting, where lower doses might be expected ^62^. Our results are also partly in contrast to Haijen et al 2018 ^45^ who described that less experienced users showed greater improvements in well-being. Nevertheless, this is a very important finding in the context of potential therapeutic use of psilocybin, as it shows that the effects of psilocybin in a controlled setting do not diminish with repeated administration, providing a rationale for repeated use in a therapeutic context.

The fact that we did not find an effect of sex on acute effects is consistent with previous work by ^44^. However, it is important to note that women in our study generally used lower doses of psilocybin than men (approximately 3 mg less) based on their weight, yet still achieved comparable intensities of psychedelic experience. This is interesting in the context of the work of Garcia-Romeu et al 2021 ^60^ who showed no significant effect of dose-adjusted effects including weight and sex variables. Also, women in our protocol were not dosed during the menstrual cycle so we partially eliminated response fluctuations because of the different phases of cycle, which has been shown in animals to interact with psilocin effects ^51^.

As mentioned in the Methods section, we decided at the time of designing the study protocol that the EEG arm would precede the fMRI arm for safety reasons. In support of this, Studerus ^44^ has also described that being placed in a PET scanner while intoxicated is associated with negative subjective experiences. It is true that not all subjects decided to proceed to the fMRI, at least some of them changed their minds and reported that they were afraid of being locked in the MRI tunnel during the experience. On the other hand, our data showed that among those who underwent all sessions, there was no significant difference in ASCs between the two settings, including the DED scale. It is worth noting that the neuroimaging nature of the study and the series of neurocognitive tasks may have prevented subjects from fully enjoying the experience. Thus, the phenomenology in our study may differ from what people might experience in a naturalistic or therapeutic setting, where the experience is usually uninterrupted by such disturbances ^63^.

Interestingly, Studerus and Haijen ^44,45^ showed that young age was also associated with negative subjective experiences. We did not observe such an effect, partly because the subjects were carefully selected as healthy volunteers and the minimum age allowed by the protocol was 28 years, with a mean age of 35.8 years.

A detailed examination of the emotional valence of the experience using the VAS scale showed, in contrast to the ASCs, a variance in the dynamics of the effects, with some subjects having experiences that oscillated between positive and negative emotional states, and others having only positive experiences. It is noteworthy that out of almost 70 experiences, only one was negative throughout the session and could be described as an overall difficult experience. On the other hand, many of the subjects actually had only positive experiences in both conditions, and at the same time almost all of them ended up in a positive mood state at the end of the session. We interpret this finding that all sessions were well guided, even in a relatively difficult neuroimaging setting ^44^ with many demanding and disturbing tasks such as time in fMRI. At the same time, it shows that the ASCs is definitely a very general scale that is not designed to capture the dynamics of the experience.

Positive persisting changes in our study were consistent but less pronounced than in previously reported studies ^5,6^. This could be due to several reasons, such as the slightly lower dose in our study, the effect of the more demanding setting due to the EEG and fMRI experiments, the blood sampled from an i.v. cannula and the neuropsychological tests in our study. It is notable that despite these differences, many of our participants (33%), as in Griffith’s study, rated the psilocybin experience as one of the top five most meaningful experiences of their lives, as the single most spiritually significant experience of their lives (15%), and as one of the top five most spiritually significant experiences (21%).

### 5.2. Relationship of subjective effects of the experience (ASCs and VAS) with persisting effects (PEQ)

Our results confirmed that OBN, VRS and GASC are significantly correlated with almost all positive subscales of PEQ, which is in line with other studies ^21,38,58^. However, we also found positive correlations of the DED with the PEQ, which contradicts previous findings that a negative/anxious part of the experience is associated with a lack of positive/antidepressant effects ^21^. However, it is important to recognise that the ASCs questionnaire is a retrospective assessment of the whole experience, neglecting the dynamics and characteristic fluctuations of the state, where DED-like experiences can switch to OBN and vice versa, and such oscillations of emotional state can occur several times during a session. As mentioned above, based on our VAS, we saw that some subjects had an overall positive experience, while others oscillated between positive and negative emotional states. In our EEG arm, it appeared that subjects with both positive and negative experiences had overall more intense experiences based on ASCs and also more positive long-term outcomes. However, as this effect was not confirmed in the fMRI arm, putting these two factors together again makes it more likely that the intensity of the experience is the main factor at play, in line with other findings ^30,32,35,36^. We hypothesized that aside from intensity, the end state of the experience could also influence the long-term effects. Our study found that the combination of peak experience and end-state positivity weakly correlated with lasting positive effects. Further exploration is needed to determine if individuals who undergo difficult breakthroughs during the session, have a peak and end up in a positive emotional state, can continue in that positive afterglow and vice versa. This could potentially shed light on why psychedelic experiences sometimes lead to persistent anxiety or negative effects in non-supportive environments. This underscores the importance of a safe and supportive setting ^64–66^.

#### 5.2.1. Psilocybin non-responders, placebo responders, nocebo effect

To our knowledge, no study to date has described details of non-responders and placebo responders to the acute effects of psilocybin. A closer look at non- or mild responders and placebo responders in our sample revealed differences in serum levels of psilocin in some, but not all, which could explain the variability. While the placebo effect has been described quite frequently, the non-response to psychedelic doses of psilocybin came as quite a surprise to us.

Regarding the serum levels of psilocin, it is worth noting that all subjects were asked to remain food free since dinner, but we did not control for what they ate and some foods could theoretically interact for longer. Furthermore we do not know much about other factors such as the microbiome, fast and slow metabolisers, etc., which could also contribute. Interestingly, on of the low responders who felt nothing in the two sessions despite having adequate psilocybin serum levels points to an individual who most likely has a lower sensitivity to psilocybin.

In our sample of 40 initially enrolled subjects, 10% appeared to be placebo responders (see online supplement). Interestingly, two subjects in the EEG group reported a placebo effect on ASCs at the second session. This is difficult to explain as the unblinding effect after the first active session was typical for most subjects. It could theoretically reflect the fact that the subjects were listening to music or sounds during the EEG arm, which could lead to scoring on the auditory questions resulting in an increase in the VRS of ASCs, or the fact that the subject felt relaxed and in a positive mood throughout the session, which could feed the OBE. Another interesting finding can be seen in **Fig. 2C**, where it is shown that in two placebo responders, the effects of ASCs are driving the improvement in long-term positive outcomes. Although we cannot statistically substantiate this observation, one can speculate that the psychedelic experience itself, regardless of the mechanism by which it was induced, could be driving long-term positive effects.

Finally, we did not observe any nocebo effect. Our sample consisted of healthy volunteers, and given the fact that psilocybin generally causes minimal serious adverse effects (apart from the difficult psychedelic experience, which is actually debatable as to whether it is an adverse effect), would be unlikely to detect such an effect.

### 5.3. Limitations

A common limitation of psychedelic trials is the process of blinding and unblinding ^67^. At the time the study was designed, it adhered to the standards of placebo-controlled trials, something that would be addressed today with an active placebo, or different doses of drug, or collection of unblinding data in a questionnaire. However, given our results showing that naivety and repeated dosing did not affect acute or long-term effects, our results are minimally biased and are in agreement with the results of Goodwin et al 2022 ^1^ where naïve participants were unable to subjectively discriminate between doses, but a dose-dependent response was still observed.

An obvious limitation of our study is the lower statistical power within the fMRI arm, where we did not expect so many people to withdraw from this part of the study. However, it should be noted that the results presented here are mostly exploratory and the study was not powered for them.

Finally, an important factor arising from non-responders and placebo/nocebo responders is that data in small sample studies can be affected by these subjects. Placebo responders had low scores for both acute and persistent effects. While this was not a major problem when comparing the whole sample, it could be a problem when we split the sample in the VAS analysis. Our sample was too small to study these two groups separately, which would probably be the best solution. Therefore, in future larger trials, non-responders and placebo/nocebo responders should also be given more attention and studied independently, as this may open up new ways of identifying them before the start of the trial/treatment.

## 6. Conclusions and implications

This study reinforces existing literature by demonstrating that a healthy, non-clinical cohort can derive substantial benefits from psilocybin experiences. These benefits include enduring positive shifts in attitudes towards self and the environment, enhanced personal meaning, spiritual significance, and overall well-being. Notably, these effects persisted following a second exposure, despite an inter-dosing interval of approximately 1.5 years. Moments of anxiety and challenge, as indicated by the Dread of Ego Dissolution (DED) scale and the Visual Analogue Scale (VAS), appear to be integral components of the psychedelic experience. Such challenging experiences do not invariably lead to negative long-term outcomes; in fact, they may contribute positively in a controlled setting. This suggests that anxiety, as a facet of the psychedelic process, should not be excessively mitigated. The depth and significance of the experience are influenced by the set and setting, which could explain the relatively lower Persisting Effects Questionnaire (PEQ) scores in our neuroimaging study compared to others. Importantly, our findings carry clinical implications, bolstering the potential of repeated psilocybin administrations in psychedelic-assisted therapy. This benefit seems to extend to patients regardless of their prior experience with psychedelics or sex.

## Data Availability

All data produced in the present study are available upon reasonable request to the authors

## Acknowledgements

This work was supported by grant from Czech Health Research Council (project NU21- 04-00307), Czech Science Foundation (projects 20-25349S), Ministry of the interior of the Czech Republic (project VK01010212), Long-term conceptual development of research organization (RVO 00023752), and Specific University Research, Czech Ministry of Education, Youth and Sports (project 260648/SVV/2024), ERDF-Project Brain dynamics, No. CZ.02.01.01/00/22_008/0004643, project VVI CZECRIN (LM2023049) and Charles University research program Cooperatio-Neurosciences and private funds obtained via PSYRES, Psychedelic Research Foundation (https://psyresfoundation.eu).

