## Supplementary material for "The phenomenology of psilocybin’s experience mediates subsequent persistent psychological effects independently of sex, previous experience or setting": Online Supplemental

**Online Supplement**

| **Inclusion Criteria** |
| --- |
| a) Men and women ages 28 to 65 |
| b) Healthy volunteers with no psychiatric history (mental illnesses included in ICD 10 F0.X - F99.X) |
| c) No family history of psychotic disorders up to second-degree relatives |

| **Exclusion Criteria:** |
| --- |
| a) Pregnancy |
| b) Intracranial hypertension, arterial hypertension or pulmonary hypertension |
| c) Stroke |
| d) Congestive heart failure |
| e) Pacemaker |
| f) Metal clamp in the head and/or face |
| g) Celiac disease |
| h) Left-handedness |
| i) Usage of any medication on daily basis except for contraception |
| j) Person in dependent position – students of medical faculties |
| k) Person in dependent position – students of other faculties younger than 28 |
| Criteria for premature termination of participation in this study: |
| a) Participant can cancel his participation anytime without giving any reason |
| b) Changes in health state including use of pharmacological substances |
| c) Changes in psychic state including traumatic events last 3 months (eg. death in family etc.) |
| d) Pregnancy |

e) Significant side effects that are related to assessments in this study

**Altered state of consciousness scale (ASCs)**

| | | DED | OBN | VRS | G-ASC |
|---|---|---|---|---|---|
| EEG | psilocybin | 28.7 (14.1) | 58.9 (23.8) | 60.1 (20.9) | 45,3 (19) |
| | placebo | 2.89 (5.32) | 6.05 (11.6) | 3 (8.08) | 3.95 (7.38) |
| | F (1,37) | 142.86 | 170.32 | 245.25 | 271.09 |
| fMRI | psilocybin | 29.1 (16.3) | 55.6 (27.5) | 60.9 (23.9) | 44.8 (17.5) |
| | placebo | 1.74 (3.27) | 2.49 (4.08) | 1.44 (2.78) | 1,89 (3) |
| | F (1,24) | 59.81 | 88.76 | 185.36 | 132.01 |

**Table 2:** ANOVA showed a significant main effect of treatment for each subscale of ASCs (mean, SD, and F values above) in both study arms. There were no significant effects of between-subjects variables (sex, experience) or interactions.

**Persisting effects Questionaire (PEQ)**

| | | Attitudes about life | Attitudes about self | Mood changes | Behavior changes | Altruistic/positive social effects | Increased spirituality |
|---|---|---|---|---|---|---|---|
| EEG | psilocybin | 39.1 (22.2) | 34.5 (21.9) | 32.3 (23.6) | 27.2 (21.2) | 31.3 (25.5) | 28.2 (24.1) |
| | placebo | 10.7 (14.9) | 9.6 (15.9) | 7.52 (11.9) | 7.24 (13.3) | 6.15 (17.3) | 8.79 (19) |
| | F (1,37) | 55.31 | 46.62 | 41.78 | 24.91 | 39.30 | 28.7 |
| fMRI | psilocybin | 30.9 (24.4) | 26.4 (22.8) | 25.3 (24.3) | 22.7 (21.6) | 24.6 (26.1) | 24.8 (25) |
| | placebo | 8.52 (19.1) | 7.92 (16.8) | 8.02 (18.6) | 7.38 (16.3) | 9.29 (22.1) | 8.25 (18.6) |
| | F (1,24) | 16.04 | 12.97 | 10.71 | 8.94 | 13.05 | 8.99 |

**Table 3:** ANOVA showed a significant main effect of treatment for each positive subscale of PEQ (mean, SD, and F values above) in both study arms. There were no significant effects of between-subjects variables (sex, experience) or interactions. We did not observe any difference in negative subscales.

### Placebo responders and psilocybin non-responders, detailed characteristics of individuals

| | Sex (M/F) | age range | weigh (kg) | dose (mg) | experienced (E) /naive (N) | mental healthcare proffesional (M) / other (O) | EEG/fMRI | session | G-ASC | PEQ positive average | Psilocin plasma levels (ng/ml) 60 min | 120 min | 240 min | 360 min |
|---|---|---|---|---|---|---|---|---|---|---|---|---|---|---|
| **PLA responders** | **M** | **31-35** | **75** | **19** | **E** | **O** | **EEG** | **1st** | **33.38** | **25.82** | n.d. | n.d. | n.d. | n.d. |
| | **F** | **36-40** | **56** | **16** | **N** | **O** | **EEG** | **1st** | **13.24** | **10.1** | n.d. | n.d. | n.d. | n.d. |
| | **F** | **26-30** | **67** | **18** | **N** | **O** | **EEG** | **2nd** | **32.04** | **38.99** | n.d. | n.d. | n.d. | n.d. |
| | **F** | **26-30** | **65** | **17** | **N** | **M** | **EEG** | **2nd** | **7.89** | **16.65** | n.d. | n.d. | n.d. | n.d. |
| **PSI non-responder or low responder** | **F** | **46-50** | **67** | **17** | **N** | **O** | **EEG** | **1st** | **0** | **0.26** | *NA* | 23 | 26 | 17 |
| | **F** | **26-30** | **60** | **16** | **E** | **M** | **EEG** | **1st** | **17.96** | **57.34** | 10 | 12 | 10 | 6 |
| | **M** | **26-30** | **72** | **19** | **E** | **O** | **EEG** | **2nd** | **12.18** | **0** | *NA* | *NA* | *NA* | *NA* |
| | **M *** | **36-40** | **77** | **20** | **E** | **M** | **EEG** | **2nd** | **20.72** | **1.03** | 24 | 25 | 14 | 7 |
| | **M** | **31-35** | **56** | **16** | **E** | **O** | **fMRI** | **1st** | **19.15** | **21.18** | *NA* | *NA* | *NA* | *NA* |
| | **M *** | **36-40** | **79** | **20** | **E** | **M** | **fMRI** | **1st** | **20.1** | **11.05** | 28 | 12 | 5 | 3 |

** identical subject*

**Table 4.** Characteristics of placebo responders and psilocybin non-responders or mild responders based on ASCs. Placebo respondent's scores equal to the psilocybin group average and/or (scores above Q3 + 1.5 * IQR) on all ASCs scales. Non-responders scores equal to placebo average and/or scores below Q1 - IQR on 3 out of 4 ASCs scales. PEQ positive average is an average of positive items on the PEQ scale. *n.d. = not detected, NA = not available.*

**Visualisation of VAS scale on emotional valence with psilocin levels and Peak + last point and correlations**

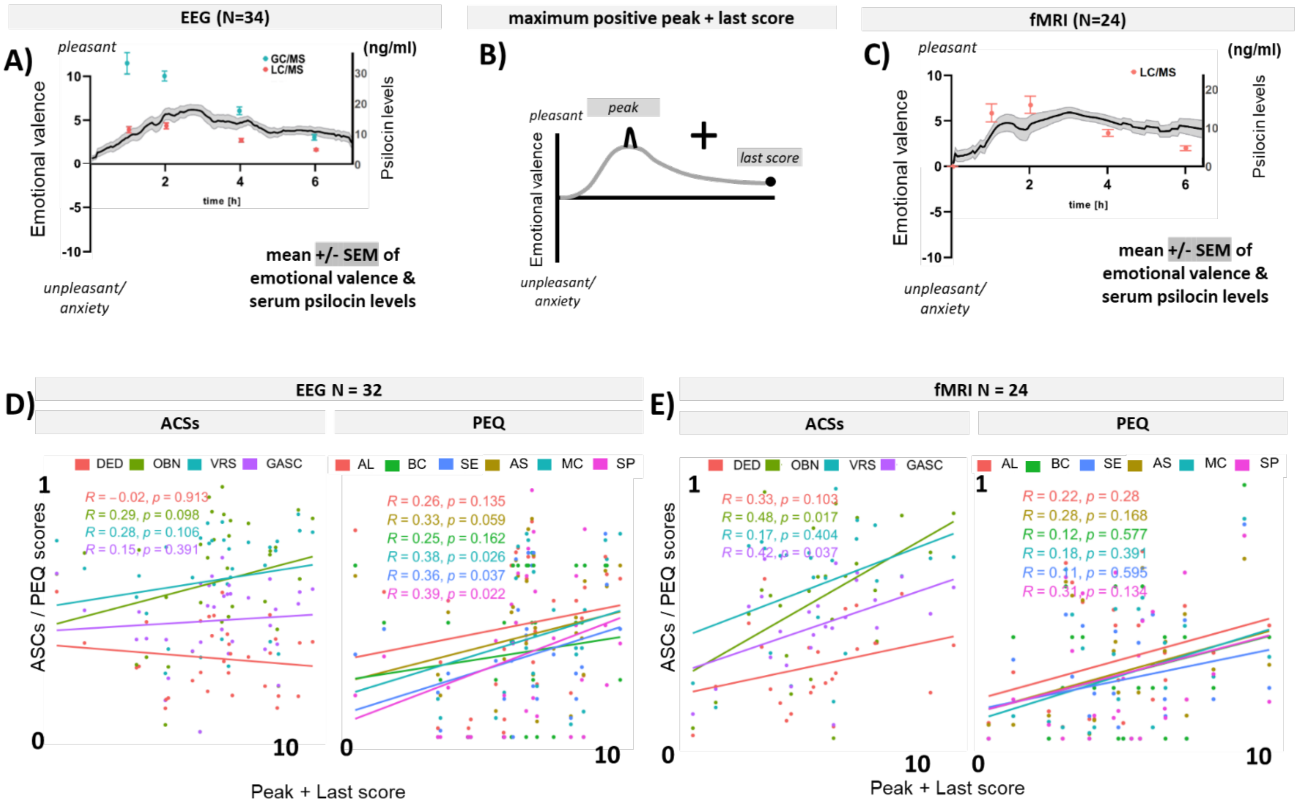


**Figure 5.** (**A, C**) visualisation of VAS scale on emotional valence with psilocin levels measured by (GC/MS and/or LC/MS) in EEG and fMRI arms. (**B**) schematic visualisation of peak + last point hypothesis. Below are correlations of ASCs and PEQ subscales with the „maximum positive peak + last score“ within EEG arm (**D**) and fMRI arm (**E**). *Shown P values are uncorrected)*
